# Background rates of hospitalizations and emergency department visits for selected thromboembolic and coagulation disorders in Ontario, Canada, 2015 to 2020, to inform COVID-19 vaccine safety surveillance

**DOI:** 10.1101/2021.04.02.21254856

**Authors:** Sharifa Nasreen, Andrew Calzavara, Maria Sundaram, Shannon E. MacDonald, Christiaan Righolt, Menaka Pai, Thalia Field, Lily W. Zhou, Sarah Wilson, Jeffrey C. Kwong, on behalf of the Canadian Immunization Research Network (CIRN) Provincial Collaborative Network (PCN) investigators

**Affiliations:** Dalla Lana School of Public Health, University of Toronto, Toronto, ON, Canada; ICES, Toronto, ON, Canada; Faculty of Nursing, University of Alberta, Edmonton, AB, Canada; Vaccine and Drug Evaluation Centre, Department of Community Health Sciences, University of Manitoba, Winnipeg, MB, Canada; Department of Medicine, McMaster University, Hamilton, ON, Canada; Division of Neurology, University of British Columbia, Vancouver Stroke Program, Vancouver, BC, Canada; Stanford Stroke Center, Palo Alto, CA, USA; Public Health Ontario, ON, Canada; Department of Family and Community Medicine, University of Toronto, Toronto, ON, Canada; University Health Network, Toronto, ON, Canada

## Abstract

**Objective:** The objective of this study was to estimate background rates of selected thromboembolic and coagulation disorders in Ontario, Canada.

**Design:** Population-based retrospective observational study using linked health administrative databases. Records of hospitalizations and emergency department visits were searched to identify cases using diagnostic codes from the *International Statistical Classification of Diseases and Related Health Problems, Tenth Revision, Canada (ICD-10-CA)*.

**Participants:** All Ontario residents.

**Primary outcome measures:** Incidence rates of stroke, deep vein thrombosis, pulmonary embolism, idiopathic thrombocytopenia, disseminated intravascular coagulation, and cerebral venous thrombosis during five pre-pandemic years (2015–2019, annually, averaged, and monthly average) and 2020.

**Results:** The average annual population was 14 million with 51% female. The mean annual rates during 2015–2019 were 127.1/100,000 population (95% confidence interval [CI], 126.2, 127.9) for ischemic stroke, 22.0/100,000 (95%CI, 21.6, 22.3) for intracerebral haemorrhage, 9.4 (95%CI, 9.2, 9.7) for subarachnoid haemorrhage, 86.8/100,000 (95%CI, 86.1, 87.5) for deep vein thrombosis, 63.7/100,000 (95%CI, 63.1, 64.3) for pulmonary embolism, 6.1/100,000 (95%CI, 5.9, 6.3) for idiopathic thrombocytopenia, 1.6/100,000 (95%CI, 1.5, 1.7) for disseminated intravascular coagulation, and 1.5/100,000 (95%CI, 1.4, 1.6) for cerebral venous thrombosis. Rates were lower in 2020 than during the pre-pandemic years for ischemic stroke, deep vein thrombosis, and idiopathic thrombocytopenia. Rates were generally consistent over time, except for pulmonary embolism, which increased from 57.1 to 68.5 per 100,000 between 2015 and 2019. Rates were higher for females than males for subarachnoid haemorrhage, pulmonary embolism, and cerebral venous thrombosis, and vice versa for ischemic stroke and intracerebral haemorrhage. Rates increased with age for most of these conditions, but idiopathic thrombocytopenia demonstrated a bimodal distribution with incidence peaks at 0–19 years and ≥60 years.

**Conclusions:** Our estimated background rates help to contextualize observed events of these potential adverse events of special interest and to detect potential safety signals related to COVID-19 vaccines.

**Strengths and limitations of this study:**

➢ Recent background rates of selected thromboembolic and coagulation disorders that are potential adverse events special interest related to COVID-19 vaccine are estimated.
➢ Background rates during five pre-pandemic (2015–2019) years and 2020 will provide context for these events to identify vaccine safety signals.
➢ We used recorded diagnostic codes in administrative data without information on clinical and/or diagnostic confirmation, and the validity of these data are imperfect, which may result in under or overestimation.

## Introduction

As COVID-19 vaccines are rapidly being licensed and used in mass immunization programs worldwide, post-marketing safety surveillance is essential to maintain vaccine confidence [1]. Recently, there have been reports of serious thrombotic events from the United Kingdom, the European Union, and Scandinavian countries related to the ChAdOx1-S (AstraZeneca) vaccine.^1^ The European Medicines Agency (EMA) has reported seven cases of disseminated intravascular coagulation and 18 cases of cerebral venous thrombosis (including nine deaths) following vaccination.^2^ These events occurred predominantly in females aged <55 years. Based on pre-COVID-19 rates of these two rare conditions, the EMA’s safety committee noted that observed events among vaccinated individuals were higher than expected for that age group (expected vs. observed within 14 days of vaccination in those aged <50 years: 1 vs. 5 for disseminated intravascular coagulation, and 1.35 vs. 12 for cerebral venous thrombosis), and they recommended close safety monitoring and further analysis.^2^ A causal relationship between these two rare events and the vaccine was neither proven nor ruled out. However, the overall number of thromboembolic events after vaccination was not deemed to be higher than expected.^2^ The EMA mandated that information on the thrombotic events be added to AstraZeneca vaccine’s summary of product characteristics and package leaflets.

This situation underscores the invaluable role of knowing background rates in vaccine safety signal assessments.^3^ Without background rates, the occurrence of adverse events in vaccinated individuals could be misinterpreted as a vaccine safety signal.^4-6^ However, background rates may vary by geography, sex, age, and over time.^4 3^

Following a risk assessment based on available data on the reported events in Europe, Health Canada described in public messaging that a combination of thrombosis and thrombocytopenia had been observed very rarely following vaccination with the AstraZeneca COVID-19 vaccine, although there was no overall increased risk of thrombosis and that the benefits of the vaccine outweigh the risks. ^7^ In response to the safety signal in Europe, Health Canada also updated the product monograph for AstraZeneca to include information on the reported rare events.^8^ In it’s first statement on AstraZeneca vaccine, Canada’s National Advisory Committee on Immunization (NACI) recommended its use in adults <65 years of age due to insufficient evidence on efficacy in older adults available at that time, although the recommendation was later to expanded to individuals aged ≥18 years. ^9^ As a result of this and other factors, there is considerable variability across Canadian provinces and territories in regards to the populations offered AstraZeneca vaccine; for example, in Ontario AstraZeneca vaccine is available to adults aged ≥60 years^10^ and in British Columbia the vaccine is available for frontline priority workers^11^.However, NACI currently does not recommend AstraZencea COVID-19 vaccine in adults aged <55 years in Canada.^12^

There is a lack of Canadian data on recent background rates of potential thromboembolic and coagulation disorder adverse events of special interest (AESI) that might inform ongoing safety surveillance of AstraZeneca’s COVID-19 vaccine and others. Therefore, we sought to estimate background rates of hospitalizations and emergency department visits for selected thromboembolic and coagulation disorders during five pre-pandemic years (2015–2019) and 2020 in Ontario, Canada.

## Methods

We conducted a population-based retrospective observational study using health administrative databases from Ontario, Canada. These datasets were linked using unique identifiers and analyzed at ICES. We searched records of hospitalizations and emergency department visits to identify cases of ischemic stroke, intracerebral haemorrhage, subarachnoid haemorrhage, deep vein thrombosis, pulmonary embolism, idiopathic thrombocytopenia, disseminated intravascular coagulation, and cerebral venous thrombosis using diagnostic codes from the *International Statistical Classification of Diseases and Related Health Problems, Tenth Revision, Canada (ICD-10-CA)*^13^ (<u>Supplemental Table S1</u>). Where available, we used codes that have been previously validated.^14-18^ We used the Canadian Institute for Health Information (CIHI) Discharge Abstract Database (DAD) and CIHI’s National Ambulatory Care Reporting System (NACRS) to identify hospitalizations and emergency department visits, respectively. We included inpatient cases where an ICD-10-CA code of interest was indicated as being present on admission and of primary relevance to the stay (as opposed to being listed as a secondary diagnosis, comorbidity, or part of the medical history). We included emergency department visits where a code of interest was present in any diagnosis field. We counted only new cases after a 365-day period without the same condition (i.e., group of codes) (<u>Supplemental Table S1</u>). Information on age and sex were obtained from Ontario’s Registered Persons Database, which contains all Ontarians registered under the Ontario Health Insurance Plan.

### Statistical analysis

For each province, we calculated annual incidence rates per 100,000 population by age and sex during each of five pre-pandemic years (2015–2019) and 2020, and also the overall mean annual incidence for 2015–2019. Similarly, we calculated annual and overall mean (for 2015–2019) incidence rates separately by sex, by age group (0–19, 20–29, 30–39, 40–49, 50–59, 60–69, 70– 79 and ≥80 years), and by sex and age group. To examine seasonality, we calculated monthly averages (per 30 days) for 2015–2019. We used Statistics Canada Census and intercensal population estimates as denominators. The 95% confidence intervals (CIs) were calculated using a gamma distribution.^19^

## Results

The study population included approximately 85 million person-years of observation from 2015 to 2020. The average annual population was 14 million, with 51% female. During 2015–2019, among the 8 AESI studied, overall rates of ischemic stroke were highest at 127.1 (95%CI, 126.2, 127.9) per 100,000, whereas rates of cerebral venous thrombosis were lowest at 1.52 (95%CI, 1.43, 1.62) per 100,000 (Table 1). Rates of pulmonary embolism and cerebral venous thrombosis were higher for females aged 0–49 years than males of the same age range (Table 1, <u>Supplemental Tables S4</u> and <u>S7</u>). In contrast, rates of ischemic stroke and intracerebral haemorrhage tended to be higher for males than females (Table 1, <u>Supplemental Table S2</u>). Rates increased with age for most of these AESI, but idiopathic thrombocytopenia demonstrated a bimodal distribution with incidence peaks at 0–19 years and ≥60 years (Table 1, <u>Supplemental Table S5</u>). Rates were generally consistent over time, with the exception of pulmonary embolism, which increased from 57.1 to 68.5 per 100,000 between 2015 and 2019 (Table 2). Rates were lower in 2020 than during the pre-pandemic years for ischemic stroke, deep vein thrombosis, and idiopathic thrombocytopenia. We noted seasonality for four AESI; intracerebral haemorrhage peaked in January and December, subarachnoid haemorrhage peaked in February and December, pulmonary embolism peaked in February and October, and deep vein thrombosis peaked in April and July (<u>Supplemental Figure 1</u>).

**Table 1:** Mean background rates of hospitalizations and emergency department visits for selected thromboembolic and coagulation disorders in Ontario from 2015 to 2019.

| Age group, years | Ischemic stroke | Intracerebral haemorrhage | Subarachnoid haemorrhage | Deep vein thrombosis | Pulmonary embolism | Idiopathic thrombocytopenia | Disseminated intravascular coagulation | Cerebral venous thrombosis |
| --- | --- | --- | --- | --- | --- | --- | --- | --- |
| **Both sexes** |  |  |  |  |  |  |  |  |
| All ages | 127.07<br>(126.24, 127.91) | 21.96<br>(21.62, 22.31) | 9.43<br>(9.20, 9.66) | 86.79<br>(86.10, 87.48) | 63.68<br>(63.09, 64.27) | 6.08<br>(5.90, 6.26) | 1.55<br>(1.46, 1.65) | 1.52<br>(1.43, 1.62) |
| 0–19 | 1.78<br>(1.57, 2.00) | 1.01<br>(0.86, 1.19) | 0.68<br>(0.56, 0.83) | 4.22<br>(3.91, 4.56) | 2.25<br>(2.02, 2.50) | 6.17<br>(5.79, 6.58) | 0.45<br>(0.35, 0.57) | 1.12<br>(0.96, 1.30) |
| 20–29 | 4.68<br>(4.27, 5.13) | 2.28<br>(1.99, 2.60) | 1.80<br>(1.55, 2.09) | 26.66<br>(25.65, 27.70) | 18.83<br>(17.98, 19.71) | 2.99<br>(2.66, 3.36) | 0.96<br>(0.77, 1.17) | 1.30<br>(1.09, 1.55) |
| 30–39 | 13.07<br>(12.35, 13.83) | 3.91<br>(3.52, 4.33) | 3.96<br>(3.57, 4.39) | 53.73<br>(52.25, 55.23) | 31.34<br>(30.21, 32.49) | 3.76<br>(3.38, 4.17) | 1.07<br>(0.87, 1.30) | 1.62<br>(1.38, 1.90) |
| 40–49 | 36.88<br>(35.66, 38.14) | 8.27<br>(7.70, 8.88) | 7.90<br>(7.34, 8.49) | 80.49<br>(78.68, 82.34) | 45.68<br>(44.32, 47.07) | 3.78<br>(3.39, 4.19) | 1.04<br>(0.84, 1.27) | 1.51<br>(1.27, 1.78) |
| 50–59 | 97.01<br>(95.13, 98.93) | 17.59<br>(16.80, 18.42) | 13.97<br>(13.26, 14.71) | 107.32<br>(105.34, 109.34) | 70.85<br>(69.24, 72.49) | 4.70<br>(4.29, 5.13) | 1.77<br>(1.53, 2.05) | 1.60<br>(1.37, 1.86) |
| 60–69 | 210.41<br>(207.28, 213.58) | 33.67<br>(32.42, 34.95) | 16.49<br>(15.62, 17.39) | 152.79<br>(150.12, 155.49) | 122.76<br>(120.37, 125.18) | 6.53<br>(5.99, 7.11) | 2.63<br>(2.29, 3.01) | 1.76<br>(1.49, 2.08) |
| 70–79 | 457.78<br>(451.84, 463.79) | 78.12<br>(75.67, 80.62) | 24.24<br>(22.88, 25.65) | 228.42<br>(224.22, 232.67) | 198.19<br>(194.28, 202.15) | 12.19<br>(11.23, 13.20) | 3.52<br>(3.02, 4.09) | 2.19<br>(1.79, 2.64) |
| ≥80 | 1127.67<br>(1115.77, 1139.65) | 181.74<br>(176.98, 186.59) | 41.59<br>(39.33, 43.95) | 344.83<br>(338.27, 351.49) | 274.19<br>(268.34, 280.13) | 23.31<br>(21.63, 25.09) | 5.32<br>(4.53, 6.20) | 2.04<br>(1.56, 2.61) |
| **Females** |  |  |  |  |  |  |  |  |
| All ages | 121.52<br>(120.38, 122.67) | 20.68<br>(20.21, 21.16) | 10.56<br>(10.23, 10.90) | 86.46<br>(85.50, 87.43) | 67.33<br>(66.48, 68.18) | 6.27<br>(6.01, 6.53) | 1.52<br>(1.40, 1.66) | 1.67<br>(1.54, 1.81) |
| 0–19 | 1.64<br>(1.36, 1.95) | 0.86<br>(0.66, 1.10) | 0.52<br>(0.37, 0.70) | 4.84<br>(4.36, 5.36) | 3.37<br>(2.97, 3.81) | 5.72<br>(5.20, 6.29) | 0.44<br>(0.30, 0.61) | 0.97<br>(0.76, 1.21) |
| 20–29 | 5.46<br>(4.82, 6.17) | 2.09<br>(1.70, 2.54) | 1.67<br>(1.32, 2.08) | 32.68<br>(31.07, 34.35) | 26.33<br>(24.89, 27.84) | 3.88<br>(3.34, 4.48) | 1.18<br>(0.89, 1.53) | 1.88<br>(1.51, 2.31) |
| 30–39 | 13.77<br>(12.74, 14.87) | 3.16<br>(2.68, 3.71) | 4.41<br>(3.83, 5.05) | 60.19<br>(58.00, 62.44) | 39.25<br>(37.48, 41.07) | 5.10<br>(4.48, 5.79) | 1.37<br>(1.06, 1.75) | 2.15<br>(1.75, 2.61) |
| 40–49 | 30.84<br>(29.28, 32.46) | 6.24<br>(5.55, 6.99) | 8.53<br>(7.72, 9.40) | 80.35<br>(77.83, 82.94) | 47.06<br>(45.13, 49.05) | 4.31<br>(3.74, 4.94) | 1.01<br>(0.74, 1.34) | 1.93<br>(1.56, 2.37) |
| 50–59 | 71.20<br>(68.93, 73.53) | 14.33<br>(13.32, 15.40) | 15.56<br>(14.51, 16.67) | 90.50<br>(87.93, 93.11) | 63.81<br>(61.66, 66.01) | 4.89<br>(4.30, 5.52) | 1.61<br>(1.28, 1.99) | 1.42<br>(1.11, 1.78) |
| 60–69 | 154.37<br>(150.65, 158.16) | 25.79<br>(24.28, 27.37) | 18.23<br>(16.96, 19.56) | 131.72<br>(128.28, 135.22) | 114.65<br>(111.45, 117.93) | 6.36<br>(5.62, 7.17) | 2.55<br>(2.09, 3.08) | 1.80<br>(1.42, 2.25) |
| 70–79 | 386.64<br>(379.18, 394.22) | 65.48<br>(62.43, 68.64) | 25.77<br>(23.87, 27.78) | 215.09<br>(209.53, 220.76) | 198.04<br>(192.71, 203.48) | 10.42<br>(9.23, 11.73) | 2.77<br>(2.17, 3.48) | 2.05<br>(1.54, 2.67) |
| ≥80 | 1111.20<br>(1096.04, 1126.53) | 174.36<br>(168.38, 180.49) | 42.22<br>(39.30, 45.29) | 343.72<br>(335.31, 352.29) | 274.15<br>(266.64, 281.82) | 20.46<br>(18.45, 22.63) | 4.23<br>(3.35, 5.28) | 2.06<br>(1.46, 2.83) |

| Males |  |  |  |  |  |  |  |  |
| --- | --- | --- | --- | --- | --- | --- | --- | --- |
| All ages | 132.78<br>(131.57, 134.00) | 23.28<br>(22.78, 23.79) | 8.27<br>(7.97, 8.57) | 87.13<br>(86.15, 88.11) | 59.93<br>(59.12, 60.75) | 5.89<br>(5.63, 6.15) | 1.58<br>(1.46, 1.72) | 1.37<br>(1.25, 1.50) |
| 0–19 | 1.91<br>(1.61, 2.24) | 1.16<br>(0.94, 1.42) | 0.85<br>(0.66, 1.07) | 3.64<br>(3.23, 4.08) | 1.19<br>(0.96, 1.45) | 6.60<br>(6.05, 7.19) | 0.47<br>(0.33, 0.64) | 1.27<br>(1.04, 1.55) |
| 20–29 | 3.96<br>(3.43, 4.55) | 2.46<br>(2.05, 2.93) | 1.93<br>(1.57, 2.35) | 21.04<br>(19.80, 22.34) | 11.82<br>(10.89, 12.80) | 2.17<br>(1.78, 2.61) | 0.75<br>(0.53, 1.03) | 0.77<br>(0.55, 1.05) |
| 30–39 | 12.36<br>(11.37, 13.41) | 4.67<br>(4.07, 5.34) | 3.51<br>(2.99, 4.09) | 47.09<br>(45.14, 49.12) | 23.22<br>(21.85, 24.65) | 2.38<br>(1.96, 2.87) | 0.76<br>(0.53, 1.05) | 1.08<br>(0.80, 1.43) |
| 40–49 | 43.19<br>(41.31, 45.14) | 10.39<br>(9.48, 11.37) | 7.24<br>(6.48, 8.06) | 80.64<br>(78.06, 83.29) | 44.25<br>(42.34, 46.22) | 3.22<br>(2.72, 3.79) | 1.07<br>(0.79, 1.42) | 1.07<br>(0.79, 1.42) |
| 50–59 | 123.18<br>(120.17, 126.25) | 20.90<br>(19.67, 22.19) | 12.35<br>(11.41, 13.35) | 124.39<br>(121.36, 127.47) | 78.00<br>(75.60, 80.45) | 4.51<br>(3.95, 5.13) | 1.94<br>(1.58, 2.36) | 1.79<br>(1.44, 2.19) |
| 60–69 | 270.67<br>(265.55, 275.86) | 42.14<br>(40.13, 44.22) | 14.61<br>(13.44, 15.86) | 175.44<br>(171.32, 179.63) | 131.47<br>(127.91, 135.10) | 6.71<br>(5.92, 7.57) | 2.72<br>(2.23, 3.29) | 1.73<br>(1.34, 2.19) |
| 70–79 | 539.40<br>(529.95, 548.98) | 92.61<br>(88.72, 96.63) | 22.48<br>(20.58, 24.50) | 243.70<br>(237.36, 250.17) | 198.35<br>(192.64, 204.19) | 14.22<br>(12.72, 15.85) | 4.39<br>(3.58, 5.34) | 2.35<br>(1.76, 3.06) |
| ≥80 | 1152.87<br>(1133.77, 1172.22) | 193.05<br>(185.27, 201.06) | 40.64<br>(37.11, 44.40) | 346.54<br>(336.10, 357.22) | 274.24<br>(264.96, 283.76) | 27.67<br>(24.78, 30.81) | 6.98<br>(5.57, 8.64) | 1.99<br>(1.28, 2.97) |

**Table 2:**
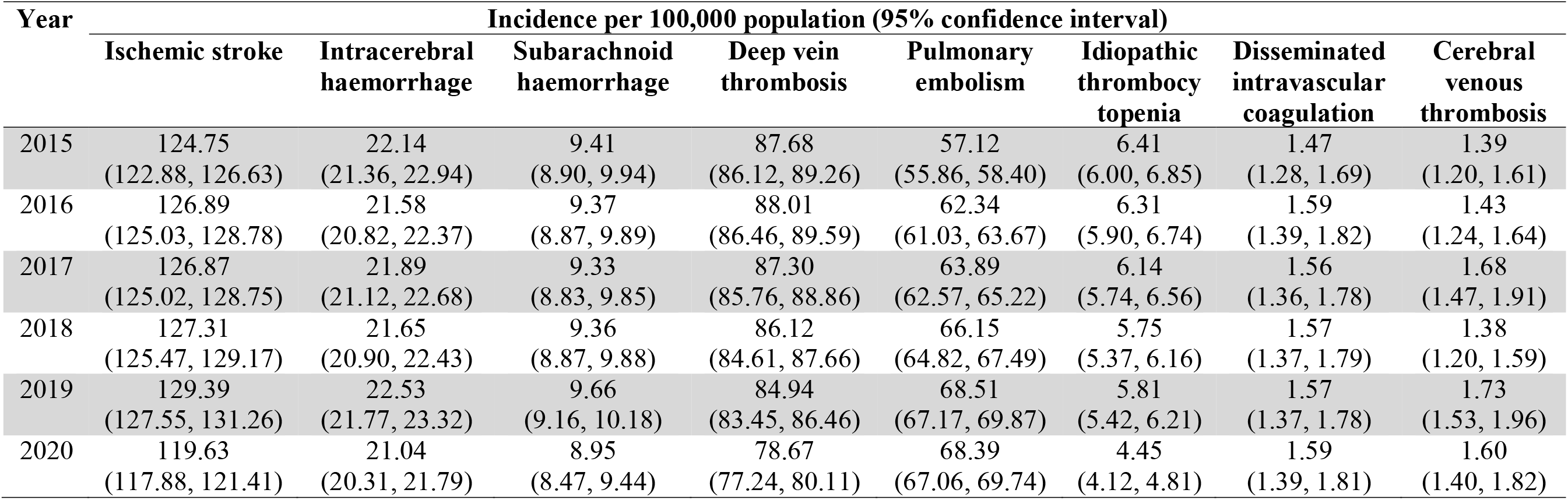
Background rates of hospitalizations and emergency department visits for selected thromboembolic and coagulation disorders for both sexes and all ages combined in Ontario, 2015 to 2020.

| Year | Incidence per 100,000 population (95% confidence interval) |  |  |  |  |  |  |  |
| --- | --- | --- | --- | --- | --- | --- | --- | --- |
|  | Ischemic stroke | Intracerebral haemorrhage | Subarachnoid haemorrhage | Deep vein thrombosis | Pulmonary embolism | Idiopathic thrombocytopenia | Disseminated intravascular coagulation | Cerebral venous thrombosis |
| 2015 | 124.75<br>(122.88, 126.63) | 22.14<br>(21.36, 22.94) | 9.41<br>(8.90, 9.94) | 87.68<br>(86.12, 89.26) | 57.12<br>(55.86, 58.40) | 6.41<br>(6.00, 6.85) | 1.47<br>(1.28, 1.69) | 1.39<br>(1.20, 1.61) |
| 2016 | 126.89<br>(125.03, 128.78) | 21.58<br>(20.82, 22.37) | 9.37<br>(8.87, 9.89) | 88.01<br>(86.46, 89.59) | 62.34<br>(61.03, 63.67) | 6.31<br>(5.90, 6.74) | 1.59<br>(1.39, 1.82) | 1.43<br>(1.24, 1.64) |
| 2017 | 126.87<br>(125.02, 128.75) | 21.89<br>(21.12, 22.68) | 9.33<br>(8.83, 9.85) | 87.30<br>(85.76, 88.86) | 63.89<br>(62.57, 65.22) | 6.14<br>(5.74, 6.56) | 1.56<br>(1.36, 1.78) | 1.68<br>(1.47, 1.91) |
| 2018 | 127.31<br>(125.47, 129.17) | 21.65<br>(20.90, 22.43) | 9.36<br>(8.87, 9.88) | 86.12<br>(84.61, 87.66) | 66.15<br>(64.82, 67.49) | 5.75<br>(5.37, 6.16) | 1.57<br>(1.37, 1.79) | 1.38<br>(1.20, 1.59) |
| 2019 | 129.39<br>(127.55, 131.26) | 22.53<br>(21.77, 23.32) | 9.66<br>(9.16, 10.18) | 84.94<br>(83.45, 86.46) | 68.51<br>(67.17, 69.87) | 5.81<br>(5.42, 6.21) | 1.57<br>(1.37, 1.78) | 1.73<br>(1.53, 1.96) |
| 2020 | 119.63<br>(117.88, 121.41) | 21.04<br>(20.31, 21.79) | 8.95<br>(8.47, 9.44) | 78.67<br>(77.24, 80.11) | 68.39<br>(67.06, 69.74) | 4.45<br>(4.12, 4.81) | 1.59<br>(1.39, 1.81) | 1.60<br>(1.40, 1.82) |

## Discussion

In this study, we estimated background rates of hospitalizations and emergency department visits for selected thromboembolic and coagulation disorders during 2015 to 2020 in Ontario. Our findings suggest that the rates were relatively stable during the pre-pandemic period but were lower in 2020 for three AESI. We observed high rates for some AESI and low rates for others. Females had higher rates of pulmonary embolism and cerebral venous thrombosis while males had higher rates of ischemic stroke and intracerebral haemorrhage.

We also observed an increasing trend in the incidence of pulmonary embolism starting before the pandemic that warrants further investigation to elucidate the cause. However, it is possible that the observed trend may have resulted from changes in clinical or coding practices over time and does not reflect a true increase in the underlying disease pattern. Lack of recent data on the trends of pulmonary embolism incidence precluded other rate comparisons.

Our estimated rates of stroke and the observed higher rates in males are comparable to previously reported global rates.^20-22^ However, higher stroke incidence was reported in females aged 18–44 years than their male counterparts (incidence rate ratio of females compared to males ranging from 1.14 to 1.93) in the Netherlands during 1998–2010.^23^ Another study in China also reported a higher age-adjusted incidence of stroke in females than males (309 vs. 280 per 100,000 population) in 2014.^24^.^25^ Our crude rates of ischemic stroke and intracerebral haemorrhage during 2015–2017 were approximately 1.5 to 2 times the age-sex standardized rates during the same period in a previous study in Ontario.^26^ However, that study included only the first-ever episode whereas we included episodes occurring after a 365-day period.

There are limitations in the estimated rates of deep vein thrombosis in recent years. Lower rates reported in older studies^27^ are likely because they included only the first incident case, whereas we included recurrent cases at least one year apart. On the other hand, the overall rate of deep vein thromboses in our study is much lower than the adult hospitalization rate reported in the USA during 2007–2009.^28^ The higher USA rates are likely because that study included multiple hospitalizations in a year, and they also included cases that developed during hospitalization from other conditions whereas our study looked at deep vein thrombosis on admission only.

There is a lack of data on the incidence of disseminated intravascular coagulation, a systemic coagulopathy that is always secondary to an underlying clinical condition, such as sepsis, malignancy, trauma, acute pancreatitis, burns, or obstetric complications.^29^ As such, studies often estimate the burden of disseminated intravascular coagulation for the underlying conditions separately. The incidence of the first episode of disseminated intravascular coagulation in ICU admitted adult patients in the USA was 18.6 per 100,000 person-years in 2010,^30^ which is far higher than the rates we have estimated.

The estimated overall incidence of cerebral venous thrombosis in our study is similar to the overall hospitalization rate of 1.75 per 100,000/year in Norway during 2011–2017^31^ and 1.32 per 100,000/year in Finland during 2005–2014.^32^ However, these rates are much higher than previously reported rates of 0.1–0.5 per 100,000 population/year.^33 18^ These differences are primarily attributable to increased use of routine vascular neuroimaging.^33^ Females in the USA had a higher incidence than males (3.02–2.69/100,000 in females vs. 0.68–1.68/100,000 in males) during 2006–2016; females aged 18–44 years also had higher incidence than their male counterparts (2.4–3.3/100,000 vs. 0.5–1.3/100,000).^34^ We also observed a higher mean incidence in females than males for all ages, and in females aged 20–49 years than their male counterparts.

Compared to pre-pandemic years, we observed lower rates in 2020 for ischemic stroke, deep vein thrombosis, and idiopathic thrombocytopenia, but not the other AESI. While thromboembolic and coagulation disorders have been associated with severe COVID-19,^35-39^ only approximately 1.3% (184,635 out of 14.7 million) of Ontarians were confirmed to be COVID-19 cases in 2020.^40^ Therefore, we would not expect COVID-19 to impact the incidence of these AESI substantially. The observed decline in the rates may have resulted from decreased healthcare-seeking observed during 2020 leading to underdiagnosis and underestimation.

Additionally, a population-level reduction in cerebrovascular events has been suggested based on a reported 12% reduction in stroke hospitalization during the peak pandemic months (March through April) in 2020 globally.^41^ This decline has been attributed to the reduced consumption of high-sodium and fast-foods, reduced exposure to ambient air pollution, improved health behaviours, and possibly from reduced exposure to other common respiratory viruses because of public health measures against COVID-19.

Our rates need to be interpreted with caution. We identified both hospitalized cases and cases that were treated in the emergency department without requiring hospitalization. Thus, our rates may be higher than those reported in the literature that used hospitalization data alone. We did not include cases that developed during the hospital stay. As a result, we may have underestimated the rates for conditions that frequently arise after hospitalization, such as disseminated intravascular coagulation and idiopathic thrombocytopenia. Additionally, we used recorded diagnostic codes in administrative data without information on clinical and/or diagnostic confirmation, and the validity of these data are imperfect, which may result in under or overestimation. However, we used previously validated codes^15-18^ or equivalent codes^14^ to improve the accuracy of case ascertainment in administrative data and there is no reason to believe that there would be substantial changes in the direction or magnitude of misdiagnosis between study years. The quality of DAD hospitalization data has been previously evaluated.^42^ Nevertheless, there may be some overlap in clinical presentations of the AESI that may have impacted our rate estimation. As background rates are population-specific, our rates may not be generalizable to other populations/settings, including jurisdictions within Canada with different population structures, distribution of risk factors, and diagnostic and coding practices.

Despite the limitations, our estimated background rates of hospitalizations and emergency department visits for the selected thromboembolic and coagulation disorders will help contextualize any observed apparent increase in these potential AESI in relation to Canada’s mass COVID-19 immunization program. This will facilitate the identification of potential safety signals and help maintain vaccine confidence by preventing misinterpretation of expected baseline event rates.

## Data Availability

The dataset from this study is held securely in coded form at ICES. While legal data sharing agreements between ICES and data providers (e.g., healthcare organizations and government) prohibit ICES from making the dataset publicly available, access may be granted to those who meet pre-specified criteria for confidential access, available at www.ices.on.ca/DAS. The full dataset creation plan and underlying analytic code are available from the authors upon request, understanding that the computer programs may rely upon coding templates or macros that are unique to ICES and are therefore either inaccessible or may require modification.

## Contributors

JCK conceived of the study design and oversaw the study. AC obtained the data and estimated incidence rates. SN drafted the manuscript. All authors interpreted the results, critically revised the manuscript, and approved the final version.

## Funding

This work was supported by the Canadian Immunization Research Network (CIRN) through a grant from the Public Health Agency of Canada and the Canadian Institutes of Health Research (CNF 151944). This study was also supported by ICES, which is funded by an annual grant from the Ontario Ministry of Health (MOH). JCK is supported by Clinician-Scientist Award from the University of Toronto Department of Family and Community Medicine. The analyses, conclusions, opinions and statements expressed herein are solely those of the authors and do not reflect those of the funding or data sources; no endorsement is intended or should be inferred.

## Conflict of interest

The authors declare no conflicts of interest.

## Patient consent for publication

Not required

## Ethics approval

ICES is a prescribed entity under Ontario’s Personal Health Information Protection Act (PHIPA). Section 45 of PHIPA authorizes ICES to collect personal health information, without consent, for the purpose of analysis or compiling statistical information with respect to the management of, evaluation or monitoring of, the allocation of resources to or planning for all or part of the health system. Projects that use data collected by ICES under section 45 of PHIPA, and use no other data, are exempt from REB review. The use of the data in this project is authorized under section 45 and approved by ICES’ Privacy and Legal Office.

## Notes

### Competing Interest Statement

The authors have declared no competing interest.

### Author Declarations

ICES is a prescribed entity under Ontarios Personal Health Information Protection Act (PHIPA). Section 45 of PHIPA authorizes ICES to collect personal health information, without consent, for the purpose of analysis or compiling statistical information with respect to the management of, evaluation or monitoring of, the allocation of resources to or planning for all or part of the health system. Projects that use data collected by ICES under section 45 of PHIPA, and use no other data, are exempt from REB review. The use of the data in this project is authorized under section 45 and approved by ICES Privacy and Legal Office.

