## Supplemental for "Background rates of hospitalizations and emergency department visits for selected thromboembolic and coagulation disorders in Ontario, Canada, 2015 to 2020, to inform COVID-19 vaccine safety surveillance"

**Supplemental material:**

### Supplemental Table S1: ICD-10-CA diagnostic codes for selected thromboembolic and coagulation disorders

| **AESI** | **Diagnosis** | **ICD-10-CA code** |
| --- | --- | --- |
| Ischemic stroke | Cerebral infarction | I63.x |
|  | Stroke, not specified as haemorrhage or infarction | I64 |
|  | Central retinal artery occlusion | H34.1 |
| Intracerebral haemorrhage | Intracerebral haemorrhage | I61.x |
| Subarachnoid haemorrhage | Subarachnoid haemorrhage | I60.x |
| Deep vein thrombosis | Phlebitis and thrombophlebitis | I80.1  I80.2  I80.3  I80.8  I80.9 |
|  | Deep phlebothrombosis in pregnancy | O22.3x |
|  | Deep phlebothrombosis in the puerperium | O87.1x |
| Pulmonary embolism | Pulmonary embolism | I26.x |
|  | Obstetric blood-clot embolism | O88.2x |
| Idiopathic thrombocytopenia (ITP) | Idiopathic thrombocytopenic purpura | D69.3x |
| Disseminated intravascular coagulation (DIC) | Disseminated intravascular coagulation | D65 |
|  | Premature separation of placenta with disseminated intravascular coagulation | O45.01x |
|  | Antepartum haemorrhage with disseminated intravascular coagulation | O46.01x |
| Cerebral venous thrombosis | Intracranial and intraspinal phlebitis and thrombophlebitis | G08 |
|  | Cerebral infarction due to cerebral venous thrombosis, non-pyogenic | I63.6 |
|  | Non-pyogenic thrombosis of intracranial venous system | I67.6 |
|  | Cerebral venous thrombosis in pregnancy | O22.5 |
|  | Cerebral venous thrombosis in the puerperium | O87.3 |

### Supplemental Table S2: Background rates of hospitalizations and emergency department visits for ischemic and haemorrhagic strokes in Ontario, 2015 to 2020

| **Age group, years** | **Incidence per 100,000 population (95% confidence interval)** | | | | | |
| --- | --- | --- | --- | --- | --- | --- |
|  | **2015** | **2016** | **2017** | **2018** | **2019** | **2020** |
| **Ischemic stroke** | | | | | | |
| **Both sexes** | | | | | | |
| All ages | 124.75 (122.88, 126.63) | 126.89 (125.03, 128.78) | 126.87 (125.02, 128.75) | 127.31 (125.47, 129.17) | 129.39 (127.55, 131.26) | 119.63 (117.88, 121.41) |
| 0–19 | 1.96 (1.50, 2.52) | 2.05 (1.57, 2.62) | 1.68 (1.25, 2.20) | 1.60 (1.19, 2.11) | 1.60 (1.19, 2.11) | 1.31 (0.94, 1.78) |
| 20–29 | 4.70 (3.77, 5.79) | 5.89 (4.85, 7.09) | 5.19 (4.22, 6.30) | 4.11 (3.28, 5.10) | 3.65 (2.88, 4.57) | 3.54 (2.79, 4.44) |
| 30–39 | 12.75 (11.15, 14.52) | 11.22 (9.74, 12.87) | 12.85 (11.28, 14.59) | 14.04 (12.41, 15.82) | 14.35 (12.73, 16.13) | 12.19 (10.72, 13.80) |
| 40–49 | 34.05 (31.47, 36.79) | 38.05 (35.31, 40.96) | 35.88 (33.21, 38.71) | 37.91 (35.16, 40.82) | 38.57 (35.79, 41.50) | 35.56 (32.89, 38.38) |
| 50–59 | 95.19 (91.03, 99.48) | 95.02 (90.88, 99.30) | 92.53 (88.44, 96.76) | 99.45 (95.20, 103.84) | 102.94 (98.61, 107.42) | 93.09 (88.95, 97.37) |
| 60–69 | 211.35 (204.17, 218.72) | 211.97 (204.89, 219.24) | 204.16 (197.29, 211.22) | 212.98 (206.04, 220.10) | 211.55 (204.73, 218.55) | 199.86 (193.32, 206.57) |
| 70–79 | 469.04 (454.96, 483.46) | 471.76 (457.89, 485.95) | 467.79 (454.39, 481.48) | 442.69 (430.00, 455.67) | 441.97 (429.57, 454.64) | 414.70 (402.93, 426.72) |
| ≥80 | 1149.95 (1122.45, 1177.94) | 1141.30 (1114.29, 1168.80) | 1135.95 (1109.33, 1163.03) | 1102.51 (1076.62, 1128.86) | 1111.62 (1085.96, 1137.72) | 986.53 (962.63, 1010.87) |
| **Females** | | | | | | |
| All ages | 118.80 (116.26, 121.39) | 121.12 (118.56, 123.71) | 122.77 (120.21, 125.37) | 121.23 (118.71, 123.80) | 123.53 (121.01, 126.10) | 111.18 (108.80, 113.60) |
| 0–19 | 1.74 (1.14, 2.55) | 1.93 (1.29, 2.77) | 1.19 (0.71, 1.88) | 1.64 (1.06, 2.42) | 1.70 (1.11, 2.49) | 1.11 (0.65, 1.78) |
| 20–29 | 6.26 (4.74, 8.11) | 6.63 (5.07, 8.52) | 5.53 (4.13, 7.25) | 5.25 (3.91, 6.90) | 3.80 (2.69, 5.22) | 3.83 (2.72, 5.24) |
| 30–39 | 13.05 (10.82, 15.60) | 11.61 (9.52, 14.02) | 12.84 (10.65, 15.34) | 14.24 (11.96, 16.83) | 16.91 (14.45, 19.68) | 12.03 (9.99, 14.36) |
| 40–49 | 30.27 (26.89, 33.97) | 30.55 (27.14, 34.27) | 32.22 (28.71, 36.03) | 30.02 (26.64, 33.72) | 31.14 (27.69, 34.90) | 30.57 (27.16, 34.29) |
| 50–59 | 72.88 (67.79, 78.25) | 65.67 (60.86, 70.77) | 70.02 (65.04, 75.28) | 69.21 (64.25, 74.45) | 78.28 (72.99, 83.85) | 62.20 (57.47, 67.20) |
| 60–69 | 151.31 (142.92, 160.07) | 155.07 (146.70, 163.79) | 154.32 (146.07, 162.93) | 157.61 (149.36, 166.20) | 153.36 (145.32, 161.72) | 145.67 (137.94, 153.72) |
| 70–79 | 389.51 (372.04, 407.58) | 392.58 (375.33, 410.41) | 399.58 (382.69, 417.01) | 377.74 (361.75, 394.26) | 376.00 (360.39, 392.10) | 341.43 (326.87, 356.47) |
| ≥80 | 1114.26 (1079.71, 1149.63) | 1141.09 (1106.50, 1176.48) | 1117.36 (1083.48, 1152.03) | 1091.55 (1058.40, 1125.47) | 1093.67 (1060.86, 1127.24) | 964.71 (934.21, 995.96) |
| **Males** | | | | | | |
| All ages | 130.88 (128.16, 133.64) | 132.84 (130.13, 135.60) | 131.09 (128.41, 133.81) | 133.54 (130.86, 136.26) | 135.40 (132.72, 138.12) | 128.29 (125.70, 130.92) |
| 0–19 | 2.17 (1.50, 3.03) | 2.16 (1.50, 3.02) | 2.15 (1.49, 3.00) | 1.57 (1.01, 2.31) | 1.50 (0.96, 2.23) | 1.50 (0.96, 2.23) |
| 20–29 | 3.22 (2.19, 4.57) | 5.20 (3.87, 6.84) | 4.87 (3.60, 6.43) | 3.06 (2.09, 4.32) | 3.52 (2.49, 4.83) | 3.28 (2.29, 4.53) |
| 30–39 | 12.44 (10.21, 15.01) | 10.82 (8.78, 13.20) | 12.87 (10.65, 15.41) | 13.84 (11.57, 16.42) | 11.78 (9.74, 14.13) | 12.35 (10.28, 14.70) |
| 40–49 | 37.96 (34.10, 42.14) | 45.86 (41.58, 50.46) | 39.70 (35.71, 44.02) | 46.19 (41.87, 50.84) | 46.38 (42.05, 51.04) | 40.82 (36.75, 45.21) |
| 50–59 | 117.79 (111.25, 124.61) | 124.71 (118.00, 131.71) | 115.33 (108.87, 122.07) | 130.12 (123.24, 137.29) | 128.01 (121.17, 135.14) | 124.57 (117.79, 131.65) |
| 60–69 | 275.97 (264.17, 288.16) | 273.30 (261.74, 285.25) | 257.84 (246.74, 269.32) | 272.43 (261.15, 284.07) | 273.89 (262.74, 285.40) | 257.74 (247.08, 268.74) |
| 70–79 | 560.85 (538.31, 584.11) | 562.69 (540.54, 585.52) | 545.95 (524.81, 567.73) | 517.07 (497.03, 537.71) | 517.47 (497.87, 537.64) | 498.60 (479.75, 518.01) |
| ≥80 | 1206.12 (1161.10, 1252.44) | 1141.63 (1098.64, 1185.87) | 1164.40 (1121.68, 1208.33) | 1119.07 (1077.88, 1161.44) | 1138.48 (1097.58, 1180.51) | 1018.95 (980.79, 1058.21) |
| **Intracerebral haemorrhage** | | | | | | |
| **Both sexes** | | | | | | |
| All ages | 22.14 (21.36, 22.94) | 21.58 (20.82, 22.37) | 21.89 (21.12, 22.68) | 21.65 (20.90, 22.43) | 22.53 (21.77, 23.32) | 21.04 (20.31, 21.79) |
| 0–19 | 0.85 (0.55, 1.24) | 1.01 (0.68, 1.43) | 0.94 (0.63, 1.35) | 1.22 (0.86, 1.67) | 1.05 (0.73, 1.48) | 0.99 (0.67, 1.41) |
| 20–29 | 2.30 (1.66, 3.09) | 2.26 (1.64, 3.05) | 2.26 (1.64, 3.03) | 2.28 (1.67, 3.04) | 2.31 (1.70, 3.06) | 1.79 (1.27, 2.46) |
| 30–39 | 3.84 (2.99, 4.86) | 3.78 (2.94, 4.78) | 3.93 (3.08, 4.94) | 3.61 (2.81, 4.57) | 4.36 (3.49, 5.39) | 4.03 (3.21, 5.00) |
| 40–49 | 8.96 (7.66, 10.42) | 8.51 (7.24, 9.94) | 7.80 (6.58, 9.18) | 7.82 (6.60, 9.20) | 8.25 (7.00, 9.67) | 8.47 (7.20, 9.90) |
| 50–59 | 17.52 (15.77, 19.42) | 17.38 (15.64, 19.27) | 15.57 (13.92, 17.37) | 18.10 (16.32, 20.03) | 19.41 (17.56, 21.41) | 19.05 (17.20, 21.04) |
| 60–69 | 32.76 (29.97, 35.74) | 32.32 (29.59, 35.24) | 36.21 (33.35, 39.25) | 31.83 (29.18, 34.65) | 35.11 (32.37, 38.03) | 31.38 (28.83, 34.11) |
| 70–79 | 86.52 (80.53, 92.84) | 76.74 (71.21, 82.59) | 80.35 (74.85, 86.14) | 74.10 (68.96, 79.52) | 74.20 (69.17, 79.50) | 70.16 (65.37, 75.21) |
| ≥80 | 187.70 (176.69, 199.22) | 185.78 (174.98, 197.08) | 177.50 (167.08, 188.41) | 179.64 (169.28, 190.47) | 178.69 (168.50, 189.35) | 160.56 (151.01, 170.56) |
| **Females** | | | | | | |
| All ages | 21.19 (20.12, 22.30) | 20.41 (19.37, 21.50) | 20.45 (19.42, 21.53) | 20.55 (19.52, 21.62) | 20.80 (19.77, 21.87) | 20.04 (19.04, 21.08) |
| 0–19 | 0.67 (0.32, 1.23) | 1.00 (0.56, 1.64) | 0.99 (0.56, 1.64) | 1.12 (0.65, 1.79) | 0.52 (0.23, 1.03) | 1.31 (0.80, 2.02) |
| 20–29 | 2.09 (1.26, 3.26) | 2.07 (1.24, 3.23) | 1.70 (0.97, 2.76) | 1.95 (1.18, 3.05) | 2.60 (1.70, 3.81) | 1.28 (0.68, 2.18) |
| 30–39 | 3.04 (2.02, 4.40) | 2.90 (1.91, 4.22) | 2.97 (1.97, 4.29) | 3.22 (2.19, 4.57) | 3.65 (2.55, 5.05) | 3.75 (2.65, 5.14) |
| 40–49 | 7.20 (5.60, 9.12) | 6.82 (5.27, 8.70) | 5.58 (4.18, 7.30) | 5.69 (4.27, 7.42) | 5.89 (4.45, 7.65) | 7.35 (5.73, 9.29) |
| 50–59 | 14.48 (12.26, 16.98) | 13.17 (11.07, 15.56) | 13.37 (11.25, 15.78) | 15.43 (13.14, 18.01) | 15.21 (12.93, 17.78) | 14.85 (12.59, 17.39) |
| 60–69 | 27.17 (23.68, 31.03) | 23.55 (20.36, 27.10) | 26.59 (23.23, 30.30) | 25.31 (22.06, 28.89) | 26.35 (23.08, 29.95) | 23.47 (20.43, 26.84) |
| 70–79 | 69.89 (62.61, 77.79) | 67.35 (60.32, 74.97) | 67.17 (60.35, 74.54) | 63.20 (56.76, 70.17) | 60.92 (54.74, 67.60) | 64.39 (58.16, 71.11) |
| ≥80 | 182.64 (168.82, 197.30) | 178.45 (164.93, 192.78) | 170.48 (157.41, 184.35) | 169.47 (156.56, 183.15) | 171.40 (158.56, 185.00) | 152.03 (140.07, 164.74) |
| **Males** | | | | | | |
| All ages | 23.12 (21.99, 24.30) | 22.79 (21.67, 23.95) | 23.37 (22.24, 24.53) | 22.78 (21.68, 23.92) | 24.30 (23.17, 25.47) | 22.06 (21.00, 23.17) |
| 0–19 | 1.02 (0.58, 1.66) | 1.02 (0.58, 1.65) | 0.88 (0.48, 1.48) | 1.32 (0.81, 2.01) | 1.56 (1.01, 2.31) | 0.69 (0.34, 1.23) |
| 20–29 | 2.50 (1.60, 3.71) | 2.45 (1.57, 3.64) | 2.78 (1.85, 4.02) | 2.58 (1.70, 3.75) | 2.04 (1.28, 3.08) | 2.27 (1.47, 3.36) |
| 30–39 | 4.68 (3.36, 6.35) | 4.69 (3.38, 6.33) | 4.91 (3.58, 6.57) | 4.01 (2.84, 5.51) | 5.08 (3.77, 6.70) | 4.31 (3.13, 5.79) |
| 40–49 | 10.78 (8.77, 13.12) | 10.26 (8.29, 12.56) | 10.12 (8.16, 12.41) | 10.06 (8.10, 12.35) | 10.74 (8.71, 13.10) | 9.65 (7.73, 11.90) |
| 50–59 | 20.60 (17.92, 23.57) | 21.64 (18.90, 24.67) | 17.80 (15.32, 20.57) | 20.81 (18.12, 23.80) | 23.68 (20.79, 26.86) | 23.33 (20.45, 26.51) |
| 60–69 | 38.77 (34.43, 43.51) | 41.78 (37.33, 46.61) | 46.58 (41.93, 51.61) | 38.83 (34.65, 43.38) | 44.50 (40.08, 49.28) | 39.83 (35.71, 44.30) |
| 70–79 | 105.72 (96.06, 116.08) | 87.53 (78.93, 96.81) | 95.45 (86.73, 104.81) | 86.59 (78.51, 95.28) | 89.40 (81.36, 98.01) | 76.77 (69.48, 84.61) |
| ≥80 | 195.67 (177.79, 214.86) | 197.14 (179.52, 216.03) | 188.25 (171.31, 206.41) | 195.02 (178.05, 213.18) | 189.62 (173.15, 207.23) | 173.24 (157.72, 189.87) |
| **Subarachnoid haemorrhage** | | | | | | |
| **Both sexes** | | | | | | |
| All ages | 9.41 (8.90, 9.94) | 9.37 (8.87, 9.89) | 9.33 (8.83, 9.85) | 9.36 (8.87, 9.88) | 9.66 (9.16, 10.18) | 8.95 (8.47, 9.44) |
| 0–19 | 0.65 (0.40, 1.01) | 0.71 (0.45, 1.08) | 0.45 (0.25, 0.76) | 0.90 (0.60, 1.30) | 0.70 (0.44, 1.06) | 0.70 (0.44, 1.06) |
| 20–29 | 1.98 (1.39, 2.72) | 2.05 (1.46, 2.81) | 1.49 (1.00, 2.14) | 1.78 (1.25, 2.47) | 1.73 (1.21, 2.40) | 1.75 (1.23, 2.41) |
| 30–39 | 4.90 (3.93, 6.04) | 3.45 (2.65, 4.41) | 4.14 (3.27, 5.18) | 3.93 (3.09, 4.92) | 3.45 (2.68, 4.37) | 3.59 (2.81, 4.51) |
| 40–49 | 8.33 (7.08, 9.74) | 8.51 (7.24, 9.94) | 6.51 (5.40, 7.78) | 7.93 (6.70, 9.32) | 8.20 (6.95, 9.61) | 7.07 (5.91, 8.39) |
| 50–59 | 14.00 (12.43, 15.71) | 14.16 (12.59, 15.88) | 13.99 (12.43, 15.69) | 13.76 (12.21, 15.45) | 13.93 (12.36, 15.64) | 13.61 (12.06, 15.31) |
| 60–69 | 14.99 (13.12, 17.05) | 16.22 (14.31, 18.32) | 17.28 (15.32, 19.42) | 15.85 (14.00, 17.88) | 17.94 (15.99, 20.05) | 14.45 (12.73, 16.33) |
| 70–79 | 24.59 (21.45, 28.07) | 24.14 (21.08, 27.52) | 23.51 (20.58, 26.73) | 23.93 (21.05, 27.10) | 24.98 (22.10, 28.13) | 23.71 (20.96, 26.72) |
| ≥80 | 42.60 (37.44, 48.27) | 40.26 (35.31, 45.70) | 43.84 (38.74, 49.43) | 40.83 (35.97, 46.16) | 40.54 (35.76, 45.78) | 38.58 (33.97, 43.63) |
| **Females** | | | | | | |
| All ages | 10.65 (9.89, 11.44) | 10.65 (9.91, 11.44) | 10.02 (9.30, 10.79) | 10.28 (9.56, 11.05) | 11.18 (10.43, 11.97) | 10.14 (9.43, 10.89) |
| 0–19 | 0.54 (0.23, 1.05) | 0.53 (0.23, 1.05) | 0.40 (0.15, 0.86) | 0.72 (0.36, 1.29) | 0.39 (0.14, 0.85) | 0.78 (0.41, 1.37) |
| 20–29 | 1.87 (1.09, 2.99) | 1.96 (1.16, 3.09) | 1.28 (0.66, 2.23) | 1.54 (0.86, 2.54) | 1.70 (0.99, 2.72) | 1.96 (1.20, 3.03) |
| 30–39 | 5.55 (4.13, 7.29) | 3.87 (2.71, 5.36) | 4.14 (2.94, 5.66) | 4.57 (3.32, 6.14) | 3.95 (2.81, 5.40) | 4.14 (2.98, 5.60) |
| 40–49 | 8.98 (7.18, 11.09) | 9.03 (7.22, 11.15) | 6.53 (5.00, 8.37) | 8.64 (6.87, 10.72) | 9.47 (7.61, 11.64) | 7.46 (5.83, 9.41) |
| 50–59 | 15.73 (13.41, 18.33) | 15.94 (13.61, 18.55) | 15.57 (13.27, 18.15) | 14.76 (12.52, 17.29) | 15.79 (13.47, 18.40) | 16.30 (13.93, 18.96) |
| 60–69 | 18.20 (15.37, 21.40) | 18.84 (16.00, 22.04) | 16.19 (13.59, 19.14) | 17.56 (14.88, 20.59) | 20.27 (17.41, 23.45) | 15.47 (13.02, 18.24) |
| 70–79 | 24.41 (20.19, 29.25) | 26.41 (22.08, 31.34) | 24.48 (20.43, 29.08) | 24.49 (20.54, 28.97) | 28.74 (24.54, 33.44) | 24.44 (20.66, 28.70) |
| ≥80 | 43.04 (36.47, 50.45) | 41.01 (34.67, 48.18) | 45.33 (38.72, 52.76) | 40.17 (34.02, 47.11) | 41.61 (35.42, 48.59) | 39.28 (33.32, 46.00) |
| **Males** | | | | | | |
| All ages | 8.14 (7.47, 8.85) | 8.05 (7.39, 8.75) | 8.62 (7.94, 9.34) | 8.42 (7.76, 9.13) | 8.10 (7.46, 8.79) | 7.72 (7.10, 8.39) |
| 0–19 | 0.77 (0.40, 1.34) | 0.89 (0.49, 1.49) | 0.51 (0.22, 1.00) | 1.07 (0.62, 1.71) | 1.00 (0.57, 1.62) | 0.62 (0.30, 1.15) |
| 20–29 | 2.08 (1.27, 3.21) | 2.14 (1.33, 3.27) | 1.69 (0.98, 2.70) | 2.01 (1.24, 3.07) | 1.76 (1.06, 2.75) | 1.55 (0.90, 2.48) |
| 30–39 | 4.22 (2.97, 5.82) | 3.01 (1.99, 4.38) | 4.14 (2.93, 5.69) | 3.27 (2.22, 4.65) | 2.95 (1.97, 4.23) | 3.04 (2.06, 4.31) |
| 40–49 | 7.66 (5.98, 9.66) | 7.97 (6.25, 10.02) | 6.49 (4.94, 8.37) | 7.18 (5.54, 9.15) | 6.86 (5.26, 8.80) | 6.65 (5.08, 8.57) |
| 50–59 | 12.25 (10.20, 14.58) | 12.37 (10.32, 14.70) | 12.38 (10.33, 14.72) | 12.74 (10.65, 15.12) | 12.04 (10.00, 14.36) | 10.88 (8.94, 13.11) |
| 60–69 | 11.54 (9.23, 14.25) | 13.40 (10.94, 16.26) | 18.45 (15.57, 21.71) | 14.02 (11.55, 16.85) | 15.44 (12.88, 18.36) | 13.36 (11.02, 16.04) |
| 70–79 | 24.80 (20.25, 30.08) | 21.54 (17.38, 26.38) | 22.39 (18.28, 27.16) | 23.30 (19.20, 28.01) | 20.68 (16.91, 25.03) | 22.88 (18.98, 27.34) |
| ≥80 | 41.90 (33.86, 51.27) | 39.09 (31.47, 47.99) | 41.56 (33.81, 50.54) | 41.82 (34.17, 50.67) | 38.94 (31.68, 47.36) | 37.53 (30.50, 45.69) |

### Supplemental Table S3: Background rates of hospitalizations and emergency department visits for deep vein thrombosis in Ontario, 2015 to 2020

| **Age group, years** | **Incidence per 100,000 population (95% confidence interval)** | | | | | |
| --- | --- | --- | --- | --- | --- | --- |
|  | **2015** | **2016** | **2017** | **2018** | **2019** | **2020** |
| **Both sexes** | | | | | | |
| All ages | 87.68 (86.12, 89.26) | 88.01 (86.46, 89.59) | 87.30 (85.76, 88.86) | 86.12 (84.61, 87.66) | 84.94 (83.45, 86.46) | 78.67 (77.24, 80.11) |
| 0–19 | 4.54 (3.82, 5.36) | 4.45 (3.74, 5.26) | 4.30 (3.60, 5.09) | 3.94 (3.28, 4.71) | 3.90 (3.24, 4.65) | 3.10 (2.51, 3.78) |
| 20–29 | 26.65 (24.37, 29.10) | 24.42 (22.24, 26.74) | 28.75 (26.42, 31.23) | 26.46 (24.26, 28.80) | 26.96 (24.78, 29.29) | 22.20 (20.24, 24.30) |
| 30–39 | 54.96 (51.59, 58.50) | 58.09 (54.65, 61.70) | 51.41 (48.21, 54.78) | 51.28 (48.12, 54.60) | 53.10 (49.93, 56.42) | 48.46 (45.48, 51.58) |
| 40–49 | 80.89 (76.88, 85.05) | 82.37 (78.31, 86.59) | 81.44 (77.38, 85.64) | 77.61 (73.65, 81.72) | 80.15 (76.13, 84.33) | 74.19 (70.32, 78.21) |
| 50–59 | 107.78 (103.36, 112.35) | 110.00 (105.54, 114.59) | 109.45 (105.00, 114.04) | 106.88 (102.48, 111.43) | 102.46 (98.13, 106.93) | 101.22 (96.90, 105.68) |
| 60–69 | 155.59 (149.43, 161.93) | 158.10 (152.00, 164.39) | 152.57 (146.64, 158.68) | 147.95 (142.18, 153.90) | 150.24 (144.50, 156.15) | 136.64 (131.24, 142.21) |
| 70–79 | 240.78 (230.72, 251.17) | 233.67 (223.94, 243.72) | 223.81 (214.58, 233.34) | 229.42 (220.31, 238.81) | 216.99 (208.33, 225.92) | 195.58 (187.53, 203.89) |
| ≥80 | 358.96 (343.67, 374.75) | 341.58 (326.88, 356.78) | 352.05 (337.30, 367.27) | 348.08 (333.60, 363.02) | 325.11 (311.30, 339.37) | 299.62 (286.52, 313.17) |
| **Females** | | | | | | |
| All ages | 87.65 (85.47, 89.88) | 87.96 (85.78, 90.18) | 87.77 (85.61, 89.97) | 85.89 (83.77, 88.05) | 83.18 (81.11, 85.29) | 76.10 (74.13, 78.11) |
| 0–19 | 5.22 (4.12, 6.51) | 5.58 (4.45, 6.91) | 5.56 (4.43, 6.88) | 3.61 (2.72, 4.70) | 4.25 (3.28, 5.42) | 3.53 (2.65, 4.60) |
| 20–29 | 33.39 (29.74, 37.36) | 28.93 (25.56, 32.62) | 36.35 (32.59, 40.41) | 32.30 (28.83, 36.08) | 32.40 (28.97, 36.13) | 25.54 (22.53, 28.84) |
| 30–39 | 62.74 (57.73, 68.08) | 64.18 (59.13, 69.54) | 56.76 (52.05, 61.78) | 58.21 (53.48, 63.23) | 59.25 (54.55, 64.25) | 53.34 (48.94, 58.03) |
| 40–49 | 80.69 (75.10, 86.59) | 82.94 (77.25, 88.93) | 80.54 (74.93, 86.45) | 80.80 (75.18, 86.73) | 76.79 (71.32, 82.57) | 72.18 (66.88, 77.78) |
| 50–59 | 90.81 (85.12, 96.79) | 95.74 (89.90, 101.85) | 91.80 (86.09, 97.79) | 87.90 (82.31, 93.78) | 86.18 (80.62, 92.01) | 80.44 (75.05, 86.10) |
| 60–69 | 133.36 (125.49, 141.60) | 139.12 (131.21, 147.40) | 132.34 (124.71, 140.33) | 126.53 (119.14, 134.25) | 127.80 (120.47, 135.45) | 113.31 (106.51, 120.44) |
| 70–79 | 224.90 (211.67, 238.74) | 213.13 (200.47, 226.37) | 222.56 (210.00, 235.67) | 215.34 (203.31, 227.90) | 201.68 (190.30, 213.56) | 180.95 (170.40, 191.99) |
| ≥80 | 360.75 (341.21, 381.12) | 344.43 (325.55, 364.13) | 347.75 (328.96, 367.33) | 351.17 (332.48, 370.64) | 316.27 (298.74, 334.56) | 299.21 (282.33, 316.83) |
| **Males** | | | | | | |
| All ages | 87.70 (85.48, 89.96) | 88.07 (85.85, 90.32) | 86.82 (84.64, 89.04) | 86.36 (84.21, 88.55) | 86.75 (84.61, 88.93) | 81.30 (79.24, 83.40) |
| 0–19 | 3.89 (2.98, 5.00) | 3.37 (2.52, 4.40) | 3.09 (2.29, 4.09) | 4.26 (3.31, 5.40) | 3.56 (2.70, 4.61) | 2.69 (1.94, 3.62) |
| 20–29 | 20.27 (17.53, 23.33) | 20.19 (17.47, 23.20) | 21.65 (18.87, 24.73) | 21.03 (18.34, 24.00) | 21.93 (19.23, 24.91) | 19.11 (16.61, 21.87) |
| 30–39 | 46.79 (42.37, 51.55) | 51.78 (47.17, 56.71) | 45.92 (41.63, 50.52) | 44.25 (40.12, 48.70) | 46.93 (42.75, 51.42) | 43.60 (39.65, 47.85) |
| 40–49 | 81.09 (75.40, 87.10) | 81.78 (76.03, 87.85) | 82.37 (76.58, 88.49) | 74.25 (68.75, 80.09) | 83.68 (77.82, 89.87) | 76.31 (70.71, 82.23) |
| 50–59 | 124.98 (118.24, 132.00) | 124.42 (117.72, 131.41) | 127.33 (120.54, 134.39) | 126.14 (119.36, 133.19) | 119.01 (112.41, 125.89) | 122.40 (115.68, 129.41) |
| 60–69 | 179.51 (170.02, 189.39) | 178.56 (169.23, 188.26) | 174.36 (165.25, 183.84) | 170.95 (162.04, 180.22) | 174.27 (165.40, 183.50) | 161.56 (153.14, 170.32) |
| 70–79 | 259.12 (243.86, 275.07) | 257.26 (242.36, 272.85) | 225.25 (211.74, 239.40) | 245.54 (231.80, 259.90) | 234.52 (221.38, 248.22) | 212.34 (200.10, 225.13) |
| ≥80 | 356.13 (331.86, 381.71) | 337.16 (313.99, 361.60) | 358.63 (335.10, 383.38) | 343.40 (320.75, 367.23) | 338.35 (316.23, 361.62) | 300.23 (279.68, 321.88) |

### Supplemental Table S4: Background rates of hospitalizations and emergency department visits for pulmonary embolism in Ontario, 2015 to 2020

| **Age group, years** | **Incidence per 100,000 population (95% confidence interval)** | | | | | |
| --- | --- | --- | --- | --- | --- | --- |
|  | **2015** | **2016** | **2017** | **2018** | **2019** | **2020** |
| **Both sexes** | | | | | | |
| All ages | 57.12 (55.86, 58.40) | 62.34 (61.03, 63.67) | 63.89 (62.57, 65.22) | 66.15 (64.82, 67.49) | 68.51 (67.17, 69.87) | 68.39 (67.06, 69.74) |
| 0–19 | 1.83 (1.38, 2.37) | 2.05 (1.57, 2.62) | 2.16 (1.68, 2.75) | 2.56 (2.03, 3.19) | 2.65 (2.11, 3.29) | 1.92 (1.46, 2.47) |
| 20–29 | 16.02 (14.26, 17.94) | 20.26 (18.29, 22.39) | 19.10 (17.21, 21.14) | 18.83 (16.98, 20.82) | 19.80 (17.94, 21.81) | 18.33 (16.55, 20.24) |
| 30–39 | 29.35 (26.89, 31.96) | 33.95 (31.33, 36.73) | 30.60 (28.14, 33.22) | 31.74 (29.27, 34.38) | 31.04 (28.63, 33.60) | 31.40 (29.02, 33.94) |
| 40–49 | 41.37 (38.52, 44.38) | 45.65 (42.64, 48.82) | 44.91 (41.92, 48.07) | 46.38 (43.33, 49.59) | 50.16 (46.99, 53.49) | 50.93 (47.73, 54.29) |
| 50–59 | 65.60 (62.16, 69.18) | 68.95 (65.43, 72.61) | 71.72 (68.12, 75.45) | 74.15 (70.49, 77.95) | 73.87 (70.21, 77.68) | 77.37 (73.60, 81.28) |
| 60–69 | 110.23 (105.06, 115.59) | 122.90 (117.52, 128.46) | 121.94 (116.64, 127.41) | 126.89 (121.55, 132.41) | 130.67 (125.32, 136.19) | 129.16 (123.92, 134.58) |
| 70–79 | 188.58 (179.69, 197.80) | 189.16 (180.41, 198.22) | 199.60 (190.88, 208.61) | 201.36 (192.83, 210.17) | 209.46 (200.95, 218.24) | 202.02 (193.83, 210.46) |
| ≥80 | 244.85 (232.25, 257.95) | 265.45 (252.50, 278.89) | 278.16 (265.07, 291.73) | 286.11 (273.00, 299.69) | 293.30 (280.20, 306.86) | 288.79 (275.93, 302.10) |
| **Females** | | | | | | |
| All ages | 60.20 (58.39, 62.05) | 65.79 (63.90, 67.71) | 66.97 (65.08, 68.89) | 70.01 (68.09, 71.96) | 73.26 (71.32, 75.24) | 70.36 (68.47, 72.29) |
| 0–19 | 2.48 (1.74, 3.41) | 3.12 (2.29, 4.15) | 3.31 (2.45, 4.36) | 4.01 (3.06, 5.14) | 3.92 (2.99, 5.05) | 2.48 (1.76, 3.41) |
| 20–29 | 23.07 (20.05, 26.41) | 27.08 (23.82, 30.66) | 26.89 (23.68, 30.41) | 25.51 (22.43, 28.89) | 28.90 (25.66, 32.43) | 25.35 (22.35, 28.63) |
| 30–39 | 35.12 (31.40, 39.17) | 43.97 (39.81, 48.44) | 37.77 (33.95, 41.91) | 39.70 (35.82, 43.89) | 39.60 (35.77, 43.73) | 41.21 (37.36, 45.36) |
| 40–49 | 42.49 (38.46, 46.82) | 46.93 (42.68, 51.49) | 45.90 (41.69, 50.42) | 48.56 (44.23, 53.21) | 51.44 (46.98, 56.21) | 53.16 (48.63, 58.00) |
| 50–59 | 60.70 (56.06, 65.62) | 62.04 (57.37, 67.00) | 64.29 (59.52, 69.34) | 64.61 (59.82, 69.68) | 67.40 (62.50, 72.59) | 66.46 (61.58, 71.63) |
| 60–69 | 102.70 (95.81, 109.96) | 116.54 (109.30, 124.13) | 112.14 (105.12, 119.51) | 116.01 (108.95, 123.42) | 124.76 (117.52, 132.32) | 115.62 (108.74, 122.81) |
| 70–79 | 187.76 (175.70, 200.44) | 181.67 (170.00, 193.93) | 202.63 (190.66, 215.16) | 205.98 (194.21, 218.27) | 208.73 (197.15, 220.82) | 194.66 (183.70, 206.09) |
| ≥80 | 242.39 (226.42, 259.19) | 266.84 (250.26, 284.25) | 269.30 (252.80, 286.59) | 292.37 (275.34, 310.18) | 297.02 (280.04, 314.76) | 282.37 (265.98, 299.51) |
| **Males** | | | | | | |
| All ages | 53.94 (52.20, 55.72) | 58.79 (56.99, 60.64) | 60.72 (58.90, 62.58) | 62.19 (60.36, 64.06) | 63.65 (61.82, 65.52) | 66.38 (64.52, 68.28) |
| 0–19 | 1.21 (0.73, 1.89) | 1.02 (0.58, 1.65) | 1.07 (0.63, 1.72) | 1.19 (0.72, 1.86) | 1.44 (0.91, 2.16) | 1.37 (0.86, 2.08) |
| 20–29 | 9.36 (7.52, 11.50) | 13.86 (11.63, 16.40) | 11.82 (9.79, 14.14) | 12.62 (10.56, 14.96) | 11.38 (9.46, 13.58) | 11.83 (9.88, 14.04) |
| 30–39 | 23.28 (20.20, 26.71) | 23.54 (20.47, 26.94) | 23.23 (20.22, 26.57) | 23.66 (20.66, 26.97) | 22.45 (19.59, 25.61) | 21.66 (18.89, 24.71) |
| 40–49 | 40.22 (36.24, 44.52) | 44.33 (40.12, 48.86) | 43.88 (39.68, 48.40) | 44.09 (39.87, 48.63) | 48.82 (44.37, 53.59) | 48.58 (44.14, 53.35) |
| 50–59 | 70.56 (65.52, 75.88) | 75.93 (70.71, 81.43) | 79.24 (73.90, 84.86) | 83.83 (78.33, 89.62) | 80.45 (75.04, 86.14) | 88.49 (82.78, 94.48) |
| 60–69 | 118.33 (110.65, 126.41) | 129.75 (121.82, 138.06) | 132.49 (124.56, 140.78) | 138.57 (130.56, 146.94) | 137.01 (129.16, 145.21) | 143.63 (135.70, 151.90) |
| 70–79 | 189.52 (176.51, 203.24) | 197.75 (184.71, 211.47) | 196.12 (183.53, 209.34) | 196.06 (183.80, 208.93) | 210.30 (197.87, 223.30) | 210.45 (198.26, 223.18) |
| ≥80 | 248.71 (228.50, 270.23) | 263.29 (242.86, 284.98) | 291.72 (270.54, 314.13) | 276.65 (256.36, 298.12) | 287.73 (267.36, 309.25) | 298.33 (277.85, 319.92) |

### Supplemental Table S5: Background rates of hospitalizations and emergency department visits for idiopathic thrombocytopenia in Ontario, 2015 to 2020

| **Age group, years** | **Incidence per 100,000 population (95% confidence interval)** | | | | | |
| --- | --- | --- | --- | --- | --- | --- |
|  | **2015** | **2016** | **2017** | **2018** | **2019** | **2020** |
| **Both sexes** | | | | | | |
| All ages | 6.41 (6.00, 6.85) | 6.31 (5.90, 6.74) | 6.14 (5.74, 6.56) | 5.75 (5.37, 6.16) | 5.81 (5.42, 6.21) | 4.45 (4.12, 4.81) |
| 0–19 | 6.56 (5.69, 7.54) | 6.11 (5.26, 7.04) | 6.82 (5.93, 7.80) | 5.67 (4.87, 6.58) | 5.72 (4.91, 6.62) | 3.35 (2.74, 4.06) |
| 20–29 | 3.90 (3.06, 4.90) | 3.05 (2.32, 3.95) | 2.88 (2.17, 3.73) | 2.38 (1.75, 3.15) | 2.84 (2.16, 3.66) | 2.65 (2.00, 3.43) |
| 30–39 | 3.51 (2.70, 4.49) | 4.05 (3.18, 5.09) | 3.93 (3.08, 4.94) | 3.77 (2.95, 4.75) | 3.55 (2.77, 4.49) | 2.75 (2.08, 3.57) |
| 40–49 | 3.61 (2.80, 4.57) | 3.91 (3.06, 4.91) | 3.60 (2.79, 4.58) | 3.94 (3.09, 4.95) | 3.83 (2.99, 4.83) | 2.48 (1.82, 3.31) |
| 50–59 | 4.73 (3.84, 5.76) | 4.95 (4.04, 6.00) | 4.95 (4.04, 6.00) | 4.30 (3.45, 5.29) | 4.56 (3.69, 5.58) | 4.26 (3.41, 5.26) |
| 60–69 | 7.75 (6.43, 9.27) | 6.58 (5.38, 7.96) | 6.31 (5.15, 7.65) | 6.16 (5.03, 7.47) | 5.94 (4.84, 7.21) | 5.10 (4.10, 6.27) |
| 70–79 | 12.97 (10.71, 15.55) | 13.58 (11.31, 16.17) | 11.75 (9.71, 14.10) | 10.28 (8.43, 12.43) | 12.58 (10.56, 14.87) | 9.52 (7.81, 11.49) |
| ≥80 | 24.24 (20.39, 28.61) | 24.93 (21.07, 29.28) | 22.17 (18.59, 26.24) | 24.66 (20.92, 28.87) | 20.74 (17.36, 24.58) | 16.47 (13.51, 19.88) |
| **Females** | | | | | | |
| All ages | 6.49 (5.91, 7.12) | 6.45 (5.87, 7.07) | 6.11 (5.55, 6.71) | 6.13 (5.57, 6.72) | 6.17 (5.61, 6.76) | 4.68 (4.20, 5.20) |
| 0–19 | 6.15 (4.96, 7.55) | 5.72 (4.57, 7.06) | 6.28 (5.08, 7.68) | 5.06 (3.99, 6.32) | 5.43 (4.32, 6.73) | 3.99 (3.05, 5.12) |
| 20–29 | 5.49 (4.08, 7.24) | 3.70 (2.56, 5.17) | 3.29 (2.24, 4.68) | 2.98 (2.00, 4.28) | 4.00 (2.86, 5.45) | 3.73 (2.64, 5.12) |
| 30–39 | 5.11 (3.76, 6.80) | 5.27 (3.90, 6.96) | 4.88 (3.57, 6.51) | 5.72 (4.31, 7.44) | 4.56 (3.32, 6.10) | 3.35 (2.32, 4.68) |
| 40–49 | 3.34 (2.28, 4.72) | 4.41 (3.18, 5.96) | 4.21 (3.01, 5.73) | 4.85 (3.55, 6.46) | 4.73 (3.45, 6.33) | 3.15 (2.13, 4.50) |
| 50–59 | 5.18 (3.89, 6.76) | 4.68 (3.46, 6.18) | 5.06 (3.79, 6.62) | 4.79 (3.56, 6.32) | 4.72 (3.49, 6.24) | 3.69 (2.61, 5.06) |
| 60–69 | 7.10 (5.38, 9.20) | 6.76 (5.11, 8.78) | 5.44 (3.98, 7.25) | 6.70 (5.09, 8.66) | 5.85 (4.37, 7.68) | 5.05 (3.69, 6.73) |
| 70–79 | 9.81 (7.20, 13.04) | 11.49 (8.70, 14.89) | 11.38 (8.69, 14.65) | 7.56 (5.45, 10.22) | 11.87 (9.24, 15.03) | 7.26 (5.28, 9.75) |
| ≥80 | 20.67 (16.20, 25.99) | 22.44 (17.82, 27.90) | 17.65 (13.62, 22.49) | 23.15 (18.54, 28.55) | 18.47 (14.42, 23.29) | 14.79 (11.23, 19.13) |
| **Males** | | | | | | |
| All ages | 6.33 (5.74, 6.96) | 6.16 (5.58, 6.78) | 6.17 (5.60, 6.78) | 5.37 (4.84, 5.93) | 5.44 (4.92, 6.01) | 4.22 (3.76, 4.72) |
| 0–19 | 6.95 (5.71, 8.39) | 6.48 (5.28, 7.86) | 7.32 (6.05, 8.79) | 6.27 (5.10, 7.62) | 6.00 (4.86, 7.32) | 2.75 (2.00, 3.69) |
| 20–29 | 2.39 (1.52, 3.59) | 2.45 (1.57, 3.64) | 2.48 (1.61, 3.67) | 1.82 (1.09, 2.84) | 1.76 (1.06, 2.75) | 1.64 (0.97, 2.59) |
| 30–39 | 1.83 (1.04, 2.97) | 2.79 (1.81, 4.12) | 2.94 (1.94, 4.28) | 1.80 (1.05, 2.87) | 2.54 (1.64, 3.75) | 2.16 (1.35, 3.26) |
| 40–49 | 3.88 (2.72, 5.37) | 3.38 (2.30, 4.80) | 2.97 (1.96, 4.32) | 2.98 (1.97, 4.34) | 2.88 (1.88, 4.22) | 1.77 (1.01, 2.88) |
| 50–59 | 4.28 (3.11, 5.74) | 5.22 (3.92, 6.81) | 4.84 (3.59, 6.38) | 3.79 (2.70, 5.18) | 4.40 (3.21, 5.89) | 4.84 (3.58, 6.40) |
| 60–69 | 8.45 (6.49, 10.81) | 6.38 (4.72, 8.43) | 7.25 (5.49, 9.40) | 5.58 (4.07, 7.47) | 6.03 (4.48, 7.95) | 5.15 (3.75, 6.92) |
| 70–79 | 16.62 (12.93, 21.03) | 15.98 (12.43, 20.22) | 12.18 (9.20, 15.81) | 13.40 (10.34, 17.08) | 13.39 (10.40, 16.97) | 12.10 (9.32, 15.45) |
| ≥80 | 29.86 (23.14, 37.93) | 28.78 (22.30, 36.55) | 29.09 (22.68, 36.75) | 26.94 (20.88, 34.21) | 24.14 (18.51, 30.95) | 18.95 (14.07, 24.99) |

### Supplemental Table S6: Background rates of hospitalizations and emergency department visits for disseminated intravascular coagulation in Ontario, 2015 to 2020

| **Age group, years** | **Incidence per 100,000 population (95% confidence interval)** | | | | | |
| --- | --- | --- | --- | --- | --- | --- |
|  | **2015** | **2016** | **2017** | **2018** | **2019** | **2020** |
| **Both sexes** | | | | | | |
| All ages | 1.47 (1.28, 1.69) | 1.59 (1.39, 1.82) | 1.56 (1.36, 1.78) | 1.57 (1.37, 1.79) | 1.57 (1.37, 1.78) | 1.59 (1.39, 1.81) |
| 0–19 | 0.62 (0.37, 0.97) | 0.36 (0.18, 0.64) | 0.55 (0.32, 0.88) | 0.45 (0.25, 0.75) | 0.29 (0.13, 0.55) | 0.51 (0.29, 0.83) |
| 20–29 | 0.69 (0.37, 1.19) | 1.05 (0.64, 1.63) | 1.23 (0.79, 1.83) | 0.89 (0.53, 1.41) | 0.91 (0.55, 1.43) | 0.76 (0.43, 1.23) |
| 30–39 | 0.95 (0.55, 1.52) | 0.77 (0.42, 1.29) | 1.40 (0.91, 2.05) | 1.15 (0.72, 1.74) | 1.07 (0.66, 1.63) | 1.28 (0.83, 1.87) |
| 40–49 | 0.90 (0.53, 1.44) | 1.34 (0.87, 1.98) | 1.08 (0.66, 1.66) | 1.19 (0.74, 1.80) | 0.70 (0.37, 1.20) | 1.46 (0.96, 2.12) |
| 50–59 | 1.88 (1.34, 2.57) | 2.21 (1.62, 2.95) | 1.15 (0.74, 1.72) | 1.88 (1.34, 2.57) | 1.75 (1.22, 2.42) | 1.62 (1.11, 2.27) |
| 60–69 | 2.52 (1.79, 3.44) | 2.69 (1.95, 3.63) | 2.33 (1.65, 3.20) | 2.45 (1.76, 3.33) | 3.14 (2.36, 4.10) | 2.15 (1.52, 2.95) |
| 70–79 | 3.58 (2.45, 5.05) | 2.80 (1.83, 4.11) | 4.46 (3.24, 5.98) | 3.08 (2.10, 4.34) | 3.67 (2.62, 5.00) | 4.05 (2.97, 5.41) |
| ≥80 | 4.50 (2.94, 6.60) | 6.06 (4.25, 8.39) | 4.43 (2.92, 6.45) | 5.92 (4.17, 8.17) | 5.61 (3.93, 7.77) | 4.88 (3.34, 6.89) |
| **Females** | | | | | | |
| All ages | 1.38 (1.12, 1.68) | 1.55 (1.27, 1.87) | 1.46 (1.19, 1.77) | 1.56 (1.29, 1.87) | 1.67 (1.39, 1.99) | 1.57 (1.30, 1.88) |
| 0–19 | 0.87 (0.46, 1.49) | 0.33 (0.11, 0.78) | 0.26 (0.07, 0.68) | 0.39 (0.14, 0.86) | 0.33 (0.11, 0.76) | 0.65 (0.31, 1.20) |
| 20–29 | 0.99 (0.45, 1.88) | 1.41 (0.75, 2.42) | 1.17 (0.58, 2.09) | 1.13 (0.56, 2.02) | 1.20 (0.62, 2.10) | 1.08 (0.54, 1.93) |
| 30–39 | 1.41 (0.75, 2.42) | 0.75 (0.30, 1.55) | 1.70 (0.97, 2.76) | 1.46 (0.80, 2.44) | 1.52 (0.85, 2.51) | 1.38 (0.75, 2.32) |
| 40–49 | 0.94 (0.43, 1.78) | 0.73 (0.30, 1.51) | 0.95 (0.43, 1.80) | 1.37 (0.73, 2.34) | 1.05 (0.50, 1.93) | 1.58 (0.88, 2.60) |
| 50–59 | 1.44 (0.81, 2.37) | 2.10 (1.32, 3.18) | 1.05 (0.52, 1.88) | 1.73 (1.02, 2.73) | 1.73 (1.03, 2.74) | 1.65 (0.96, 2.64) |
| 60–69 | 1.99 (1.14, 3.24) | 2.54 (1.57, 3.88) | 2.48 (1.54, 3.79) | 2.31 (1.41, 3.57) | 3.38 (2.28, 4.82) | 1.76 (1.00, 2.85) |
| 70–79 | 1.67 (0.72, 3.29) | 2.82 (1.54, 4.74) | 3.79 (2.32, 5.86) | 2.88 (1.65, 4.68) | 2.58 (1.44, 4.26) | 2.97 (1.76, 4.70) |
| ≥80 | 3.68 (1.96, 6.29) | 5.54 (3.39, 8.56) | 3.26 (1.68, 5.69) | 3.99 (2.23, 6.58) | 4.68 (2.77, 7.40) | 4.08 (2.33, 6.63) |
| **Males** | | | | | | |
| All ages | 1.57 (1.29, 1.90) | 1.64 (1.35, 1.97) | 1.67 (1.38, 2.01) | 1.59 (1.31, 1.91) | 1.46 (1.20, 1.77) | 1.61 (1.33, 1.93) |
| 0–19 | 0.38 (0.14, 0.83) | 0.38 (0.14, 0.83) | 0.82 (0.44, 1.40) | 0.50 (0.22, 0.99) | 0.25 (0.07, 0.64) | 0.37 (0.14, 0.82) |
| 20–29 | 0.42 (0.11, 1.06) | 0.71 (0.29, 1.47) | 1.29 (0.69, 2.21) | 0.67 (0.27, 1.38) | 0.65 (0.26, 1.33) | 0.45 (0.15, 1.06) |
| 30–39 | 0.46 (0.12, 1.17) | 0.78 (0.31, 1.61) | 1.09 (0.52, 2.01) | 0.84 (0.36, 1.66) | 0.61 (0.22, 1.33) | 1.18 (0.61, 2.05) |
| 40–49 | 0.86 (0.37, 1.70) | 1.97 (1.16, 3.11) | 1.21 (0.60, 2.16) | 0.99 (0.45, 1.89) | 0.33 (0.07, 0.97) | 1.33 (0.69, 2.32) |
| 50–59 | 2.33 (1.49, 3.47) | 2.32 (1.49, 3.45) | 1.26 (0.67, 2.15) | 2.04 (1.26, 3.12) | 1.76 (1.04, 2.78) | 1.58 (0.90, 2.57) |
| 60–69 | 3.09 (1.96, 4.63) | 2.86 (1.79, 4.33) | 2.16 (1.26, 3.46) | 2.61 (1.61, 3.98) | 2.89 (1.85, 4.31) | 2.58 (1.62, 3.90) |
| 70–79 | 5.78 (3.70, 8.60) | 2.78 (1.44, 4.85) | 5.22 (3.34, 7.76) | 3.30 (1.89, 5.36) | 4.92 (3.19, 7.27) | 5.29 (3.52, 7.65) |
| ≥80 | 5.79 (3.09, 9.91) | 6.87 (3.93, 11.16) | 6.23 (3.49, 10.28) | 8.85 (5.54, 13.39) | 7.01 (4.15, 11.08) | 6.07 (3.47, 9.85) |

### Supplemental Table S7: Background rates of hospitalizations and emergency department visits for cerebral venous thrombosis in Ontario, 2015 to 2020

| **Age group, years** | **Incidence per 100,000 population (95% confidence interval)** | | | | | |
| --- | --- | --- | --- | --- | --- | --- |
|  | **2015** | **2016** | **2017** | **2018** | **2019** | **2020** |
| **Both sexes** | | | | | | |
| All ages | 1.39 (1.20, 1.61) | 1.43 (1.24, 1.64) | 1.68 (1.47, 1.91) | 1.38 (1.20, 1.59) | 1.73 (1.53, 1.96) | 1.60 (1.40, 1.82) |
| 0–19 | 1.18 (0.82, 1.63) | 1.01 (0.68, 1.43) | 1.26 (0.90, 1.72) | 0.87 (0.57, 1.26) | 1.31 (0.94, 1.78) | 0.80 (0.52, 1.18) |
| 20–29 | 0.96 (0.57, 1.52) | 1.26 (0.81, 1.88) | 1.49 (1.00, 2.14) | 1.29 (0.84, 1.89) | 1.49 (1.01, 2.11) | 1.56 (1.07, 2.19) |
| 30–39 | 1.78 (1.22, 2.52) | 1.81 (1.24, 2.54) | 1.77 (1.22, 2.49) | 1.20 (0.76, 1.81) | 1.57 (1.07, 2.23) | 1.23 (0.80, 1.81) |
| 40–49 | 1.64 (1.12, 2.33) | 1.45 (0.95, 2.10) | 1.45 (0.96, 2.11) | 1.13 (0.70, 1.73) | 1.89 (1.31, 2.63) | 1.51 (1.00, 2.18) |
| 50–59 | 1.35 (0.90, 1.95) | 1.54 (1.05, 2.17) | 1.59 (1.09, 2.23) | 1.79 (1.26, 2.46) | 1.75 (1.22, 2.42) | 1.96 (1.40, 2.67) |
| 60–69 | 1.36 (0.84, 2.07) | 1.38 (0.86, 2.09) | 2.02 (1.39, 2.84) | 1.50 (0.97, 2.21) | 2.50 (1.81, 3.37) | 2.15 (1.52, 2.95) |
| 70–79 | 1.56 (0.86, 2.63) | 1.94 (1.15, 3.07) | 2.84 (1.89, 4.10) | 2.40 (1.55, 3.55) | 2.11 (1.34, 3.17) | 2.73 (1.86, 3.88) |
| ≥80 | 1.90 (0.95, 3.41) | 1.85 (0.92, 3.32) | 2.30 (1.26, 3.86) | 2.24 (1.23, 3.76) | 1.87 (0.97, 3.27) | 2.44 (1.39, 3.96) |
| **Females** | | | | | | |
| All ages | 1.55 (1.27, 1.87) | 1.51 (1.23, 1.82) | 1.77 (1.47, 2.10) | 1.59 (1.31, 1.90) | 1.94 (1.64, 2.29) | 1.74 (1.46, 2.07) |
| 0–19 | 0.94 (0.51, 1.57) | 0.86 (0.46, 1.48) | 0.79 (0.41, 1.39) | 1.12 (0.65, 1.79) | 1.11 (0.65, 1.78) | 0.85 (0.45, 1.45) |
| 20–29 | 1.43 (0.76, 2.44) | 1.63 (0.91, 2.69) | 2.66 (1.72, 3.92) | 1.54 (0.86, 2.54) | 2.10 (1.30, 3.21) | 2.16 (1.35, 3.27) |
| 30–39 | 2.39 (1.50, 3.62) | 2.15 (1.31, 3.32) | 2.44 (1.55, 3.66) | 1.66 (0.95, 2.70) | 2.13 (1.32, 3.25) | 2.17 (1.36, 3.28) |
| 40–49 | 1.98 (1.19, 3.10) | 1.89 (1.12, 2.99) | 1.68 (0.96, 2.74) | 1.90 (1.12, 3.00) | 2.21 (1.37, 3.38) | 1.89 (1.12, 2.99) |
| 50–59 | 1.15 (0.59, 2.01) | 1.05 (0.52, 1.88) | 1.34 (0.73, 2.24) | 1.63 (0.95, 2.61) | 1.93 (1.18, 2.97) | 1.84 (1.11, 2.88) |
| 60–69 | 1.74 (0.95, 2.93) | 1.33 (0.66, 2.38) | 1.77 (0.99, 2.92) | 1.50 (0.80, 2.57) | 2.59 (1.64, 3.89) | 1.43 (0.76, 2.44) |
| 70–79 | 1.25 (0.46, 2.72) | 1.81 (0.83, 3.44) | 2.47 (1.31, 4.22) | 2.16 (1.12, 3.77) | 2.41 (1.32, 4.04) | 2.48 (1.39, 4.08) |
| ≥80 | 2.27 (0.98, 4.46) | 2.49 (1.14, 4.73) | 2.17 (0.94, 4.28) | 1.86 (0.75, 3.84) | 1.56 (0.57, 3.40) | 2.04 (0.88, 4.02) |
| **Males** | | | | | | |
| All ages | 1.23 (0.98, 1.53) | 1.35 (1.08, 1.65) | 1.59 (1.30, 1.91) | 1.18 (0.94, 1.46) | 1.52 (1.25, 1.83) | 1.46 (1.19, 1.76) |
| 0–19 | 1.40 (0.88, 2.12) | 1.14 (0.68, 1.81) | 1.70 (1.12, 2.48) | 0.63 (0.30, 1.15) | 1.50 (0.96, 2.23) | 0.75 (0.39, 1.31) |
| 20–29 | 0.52 (0.17, 1.21) | 0.92 (0.42, 1.74) | 0.40 (0.11, 1.02) | 1.05 (0.52, 1.88) | 0.93 (0.44, 1.70) | 1.00 (0.50, 1.79) |
| 30–39 | 1.14 (0.55, 2.10) | 1.45 (0.77, 2.48) | 1.09 (0.52, 2.01) | 0.74 (0.30, 1.52) | 1.02 (0.49, 1.87) | 0.29 (0.06, 0.86) |
| 40–49 | 1.29 (0.67, 2.26) | 0.98 (0.45, 1.87) | 1.21 (0.60, 2.16) | 0.33 (0.07, 0.97) | 1.55 (0.85, 2.60) | 1.11 (0.53, 2.04) |
| 50–59 | 1.55 (0.89, 2.53) | 2.03 (1.26, 3.10) | 1.84 (1.11, 2.87) | 1.95 (1.19, 3.00) | 1.57 (0.90, 2.54) | 2.08 (1.29, 3.17) |
| 60–69 | 0.94 (0.38, 1.93) | 1.43 (0.71, 2.56) | 2.29 (1.36, 3.62) | 1.49 (0.77, 2.60) | 2.41 (1.47, 3.73) | 2.93 (1.90, 4.32) |
| 70–79 | 1.93 (0.83, 3.80) | 2.08 (0.95, 3.96) | 3.26 (1.83, 5.38) | 2.68 (1.43, 4.58) | 1.77 (0.81, 3.36) | 3.03 (1.73, 4.91) |
| ≥80 | 1.34 (0.28, 3.91) | 0.86 (0.10, 3.10) | 2.49 (0.92, 5.43) | 2.81 (1.13, 5.80) | 2.34 (0.86, 5.08) | 3.03 (1.31, 5.98) |

### Supplemental Figure 1: Monthly average incidence rates of hospitalization and emergency department visits in Ontario, 2015 to 2020: A. Stroke, B. Deep vein thrombosis, C. Pulmonary embolism, D. Idiopathic thrombocytopenia, E. Disseminated intravascular coagulation, F. Cerebral venous thrombosis

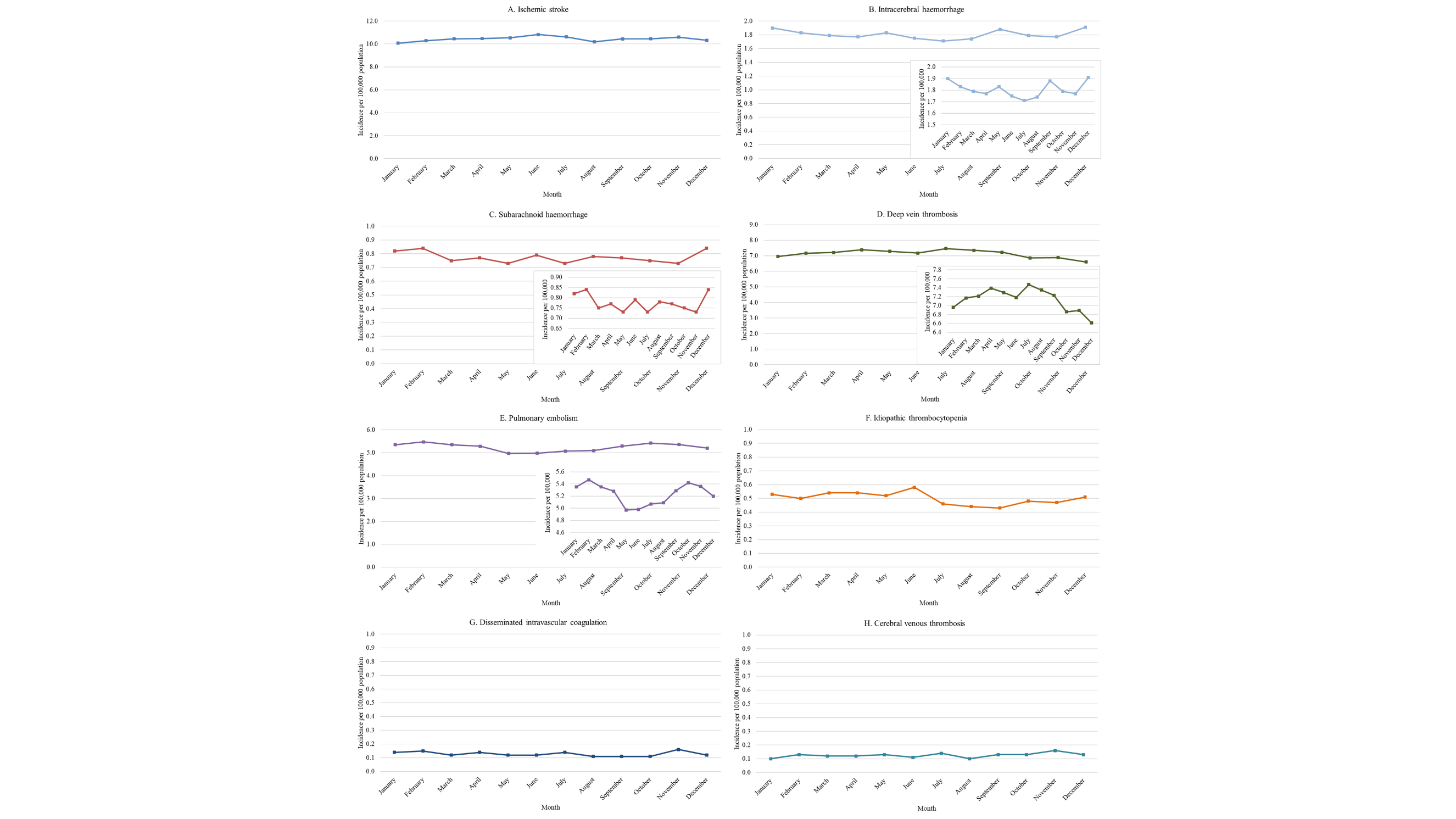
